# Safety of BNT162b2 mRNA COVID-19 vaccine batches: A nationwide cohort study

**DOI:** 10.1101/2024.01.22.24301520

**Authors:** Anders Hviid, Ingrid Bech Svalgaard

**Author notes:** Corresponding author: Anders Hviid, Department of Epidemiology Research, Statens Serum Institut, Artillerivej 5, 2300 Copenhagen S, Denmark.

## Abstract

**Background:** The safety of the BNT162b2 mRNA COVID-19 vaccine has been extensively evaluated since the global rollout began. While serious adverse events are rare, safety issues continue to arise. This study evaluates the claim that earlier small vaccine batches were associated with higher rates of serious adverse events compared to later batches.

**Methods:** A nationwide cohort study was conducted in Denmark, comprising individuals vaccinated with the BNT162b2 vaccine from 52 pre-defined batches classified into three pre-defined groups. Vaccinated individuals were matched 1:1 between batch groups on age, sex, and vaccination priority group. The study outcomes, included 27 serious adverse events, 2 negative control outcomes and all-cause mortality. Cox regression was used to estimate hazard ratios (HRs) comparing rates between batch groups in the 28-days following vaccination. We conducted two comparisons of the early small batches to two groups of larger batches used later in the pandemic.

**Results:** In the study period, 9,983,448 vaccinations were administered from batches in the three pre-defined groups. Slightly increased rates of arrhythmia were observed in both study comparisons, HRs 1.25 (95% CI,1.05-1.50) and 1.15 (1.00-1.31), respectively, but sensitivity analyses did not robustly support these associations. For the remaining outcomes, increased rates in both study comparisons were not observed.

**Conclusion:** This nationwide cohort study provides reassurance regarding the safety of the BNT162b2 vaccine across different batches used in Denmark. The findings support the overall safety of the vaccine, with no clinically relevant variations in serious adverse event rates between batches.

## Introduction

The safety of the BNT162b2 mRNA COVID-19 vaccine has been scrutinized extensively and serious adverse events are rare. However, given the vast number of doses administered globally, it is to be expected that even rare serious adverse events occur shortly after vaccination purely by chance. Many pharmacovigilance reporting systems were initially overwhelmed by the large influx of reports of adverse events, including serious adverse events, at the start of the vaccination rollouts. This was a result of the sheer scale of the rollout, the call for all adverse events to be reported and the fact that vulnerable elderly and healthcare workers were the first in line for vaccination.

A Danish group of researchers has questioned this interpretation of the initial spike in reports and has instead claimed that the earliest batches had safety issues which the later batches did not have.[1] This claim was based on data obtained through the Danish “public access to information”-act. The data in question comprised the number of doses delivered according to batch numbers and the number of adverse events reported according to batch numbers. Their conclusion was that the smaller batches delivered early in the vaccination roll-out had significantly higher rates of adverse events than larger batches delivered later in the roll-out.

To evaluate the hypothesis that the earlier batches were associated with higher rates of serious adverse events and to circumvent the limitations of pharmacovigilance reports for causal assessments, we conducted a nationwide cohort study of the association between groups of batches and 28 outcomes; 27 diagnosed in the hospital setting and all-cause mortality.

## Methods

### Study population

We designed a cohort study comprising all individuals living in Denmark and vaccinated at least once with a BNT162b2 vaccine (original monovalent version) from one of 52 pre-defined batches. The 52 batches were the same ones as those included in a previous research letter based on data obtained through the Danish “public access to information”-act.[1,2] We verified that these batches matched batches recorded in the Danish Vaccination Register.[3] The batches were classified into 3 groups according to the those previously presented in the research letter: Group 1, with almost no adverse event reports (0-0.19 reports per 1000 doses), group 2 with slightly higher reporting rates (0.61-7.70 reports per 1000 doses) and group 3 with high reporting rates (14.7-184.6 reports per 1000 doses).[1] We then matched the vaccinations in group 3 1:1 with group 1 and group 2, respectively. We used exact matching on age (10-year intervals), sex and vaccination priority group (persons living in nursing homes, persons 65 years or older, who receive certain types of home care, selected patients with conditions that carry a significant increased risk of a severe course of COVID-19, health care personnel and the general population)

### Outcomes

We included 30 study outcomes comprising 27 adverse events adapted from prioritised lists of adverse events of special interest for the covid-19 vaccines,[4] 2 negative control outcomes and all-cause mortality. Study outcomes were identified using International Classification of Disease 10^th^ revision (ICD-10) codes (supplementary table 1) assigned discharge diagnoses for hospital contacts recorded in the Danish National Patient Register.[5] All-cause mortality was identified from the Danish Civil Registration System.[6] We included primary diagnoses and inpatient, outpatient and emergency department contacts. The date of admission served as the event date.

### Statistical analyses

The matched pairs were followed for 28-days for the occurrence of the study outcomes. Each of the 30 outcomes were studied separately, meaning that outcomes not under study did not censor follow-up for the outcome under study. No individuals were eligible for matching if they had a history of the outcome in the 2-years preceding vaccination. Follow-up was censored in the case of death, emigration or disappearance from the national registers. We used Cox proportional hazards regression with time since vaccination as the underlying time-scale to estimate hazard ratios (HRs) with 95% confidence intervals (CIs) comparing the hazards of the study outcomes in group 3 vs. group 1 and group 2, respectively. Since individuals could contribute with multiple vaccinations, we used robust standard errors. We conducted sensitivity analyses in the form of age-, sex- and priority group stratification of associations that were statistically significant in both comparisons.

## Results

A total of 9,983,728 BNT162b2 vaccinations from the 3 batch groups were recorded; 2,647,879 (27%) from group 1, 6,935,006 (69%) from group 2 and 400,843 (4%) from group 3. The included vaccinations were administered from December 27, 2020 to April 25, 2023; although very few after March 2022 (figure 1). Vaccinations from group 3 were administered from December 27, 2020 to October 28, 2022(median date, January 30, 2021). Group 3 recipients were more likely to be elderly (33.4% were 80+-year-olds), female (68.2%) and had significant proportions of high-risk individuals (10.1%) and frontline personnel (40.1%) (table 1). Vaccinations from group 2 were administered from December 31, 2020 to November 11, 2022 (median date, June 26, 2021). Group 2 recipients were more likely to be middle-aged individuals (34.7% were 40-59-year-olds) (table 1). Vaccinations from group 1 were administered from January 7, 2021 to April 25, 2023 (median date, December 20, 2021). Group 1 recipients also had a high proportion of middle-aged individuals (32.6% were 40-59-year-olds) (table 1).

**Figure 1.**
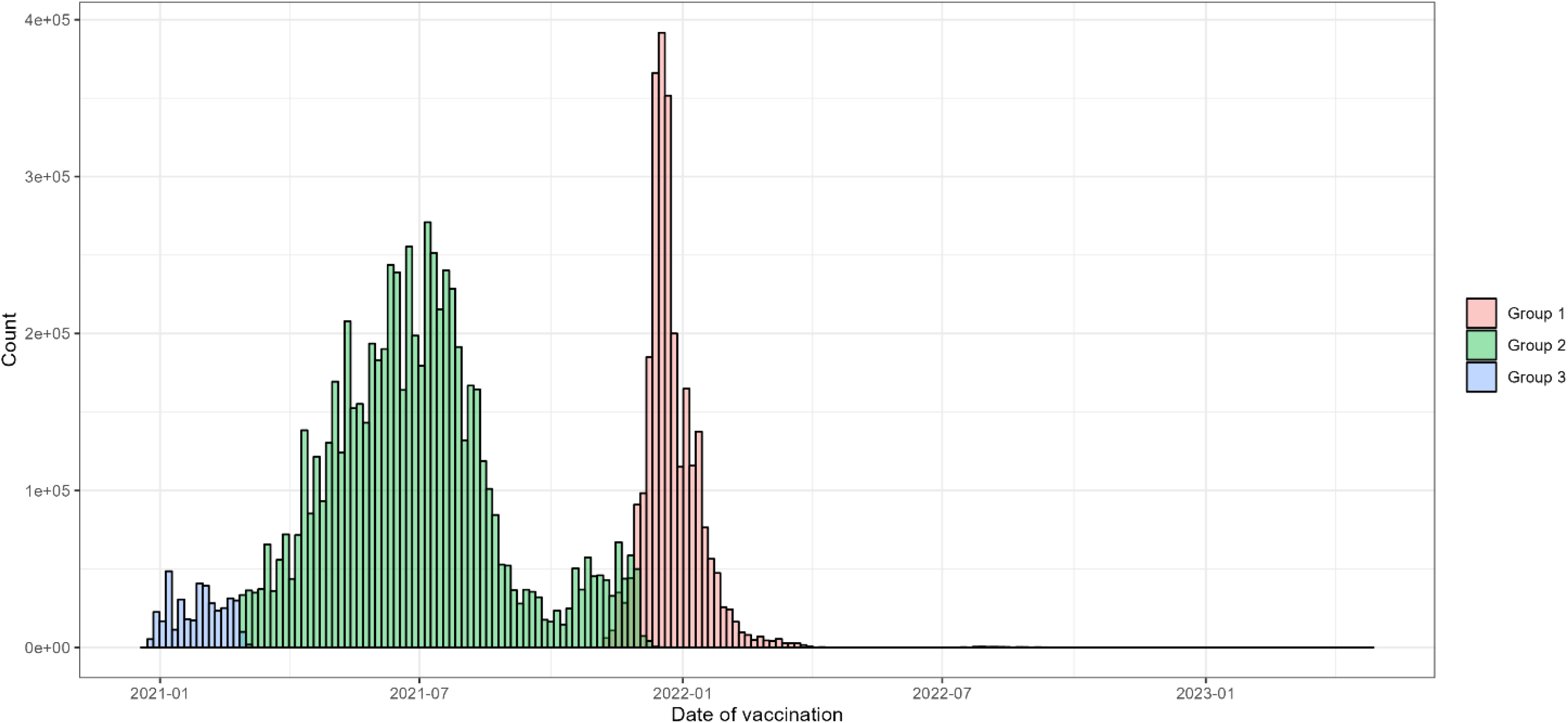
Distribution of vaccination dates for the three vaccine batch groups.

**Table 1.**
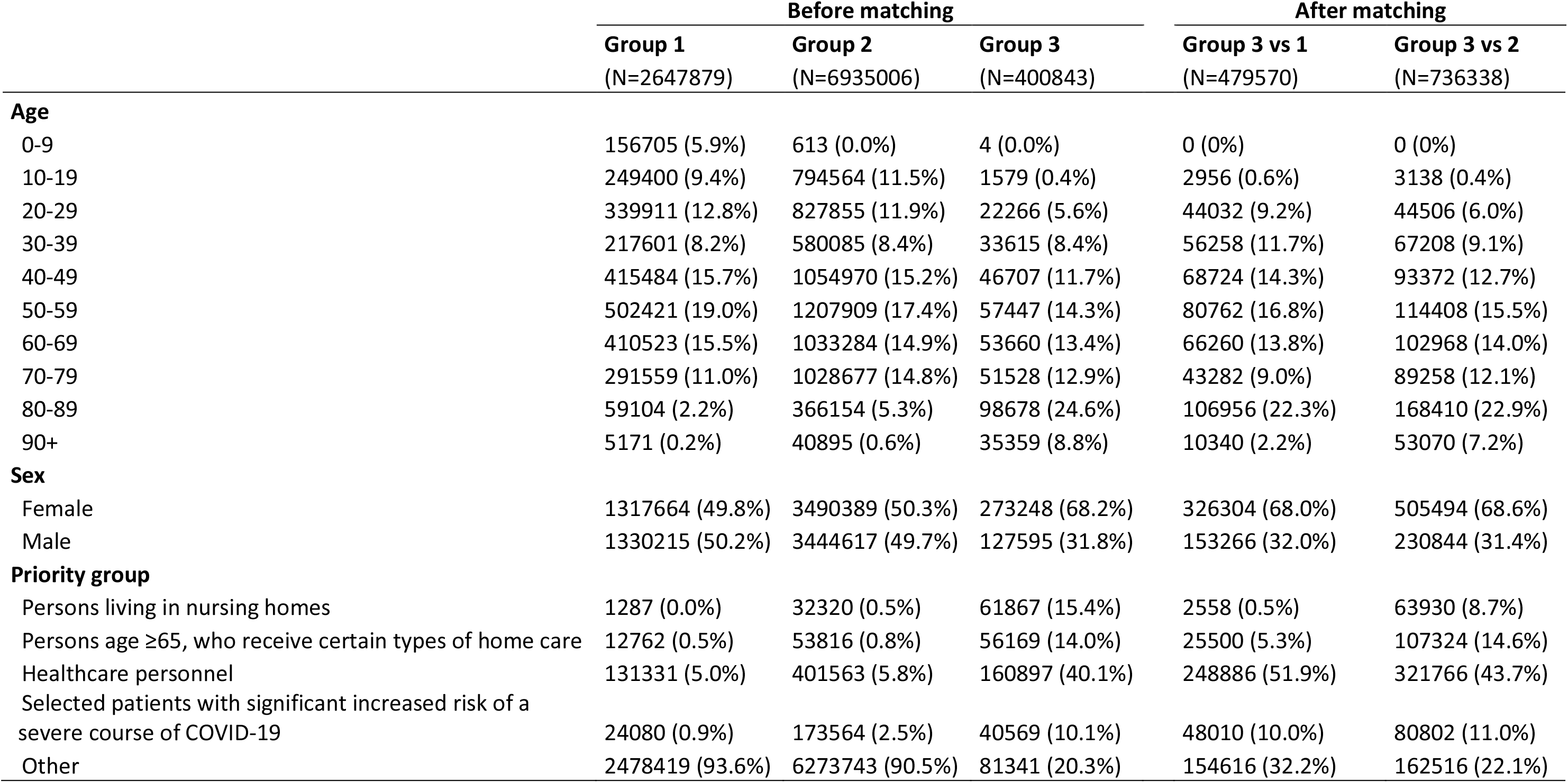
Characteristics of each person in each vaccine batch group before matching and after matching (before outcome specific exclusions).

We were able to match 239,785 pairs 1:1 in the group 3 vs 1 comparison (59.8% of group 3) and 368,169 pairs 1:1 in the group 3 vs 2 comparison (91.8% of group 3). The matched cohorts had higher proportions of elderly (^∼^22% 80-89-year-olds), females (^∼^68%), healthcare personnel (52% and 44% respectively) and high-risk individuals (^∼^10%) (table 1).

In the follow-up for study outcomes in the group 3 vs group 1 comparison, we were able to include 453,832 to 479,528 individuals (depending on the specific study outcome and the number excluded due to a history of the outcome) (figure 2). The rates of cerebrovascular- and ischemic cardiovascular events were not increased in group 3 compared to group 1, HRs 0.94 (95% CI, 0.73-1.22) and 1.01 (0.76-1.35), respectively (figure 2). Out of the 28 study outcomes, only arrythmia and thrombocytopenia and other coagulative disorders were observed at significantly higher rates in group 3, HRs 1.25 (95% CI, 1.05-1.50) and 5.25 (1.8-15.29). A number of study outcomes were very rare, and some, such as Guillain Barré syndrome and transverse myelitis, were not observed in either group 3 or group 1 (figure 2). The HRs for both of the negative control outcomes were close to 1. All-cause mortality was decreased, HR 0.81 (95% CI, 0.71-0.93).

**Figure 2.**
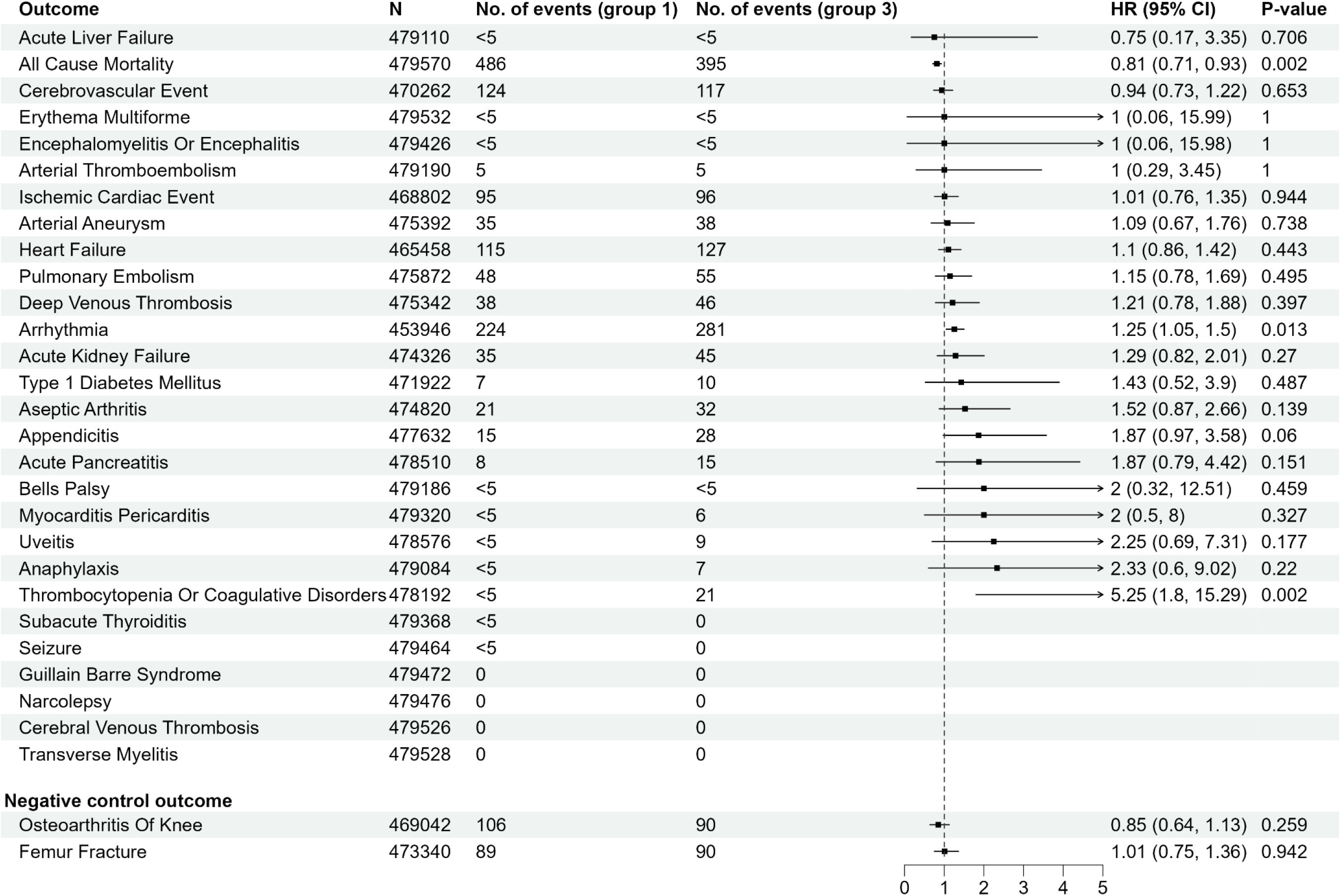
Main analysis comparing vaccine batch group 3 and vaccine batch group 1.

We were able to include 736,302 to 712,310 individuals in the group 3 vs group 2 comparison (figure 3). The rates of cerebrovascular- and ischemic cardiovascular events were not increased in group 3 compared to group 2, HRs 1.01 (95% CI,0.84-1.22) and 1.07 (0.85-1.35), respectively (figure 3). Out of the 28 study outcomes, only arrythmia, deep vein thrombosis and all-cause mortality were observed at significantly higher rates in group 3 compared to group 2, HRs 1.15 (95% CI, 1.00-1.31), 1.36 (1.00-1.85) and 1.09 (1.01-1.17), respectively. The HR for the negative control outcome osteoarthritis of the knee was reduced in group 3 compared to group 2, HR 0.74 (95% CI, 0.59-0.92).

**Figure 3.**
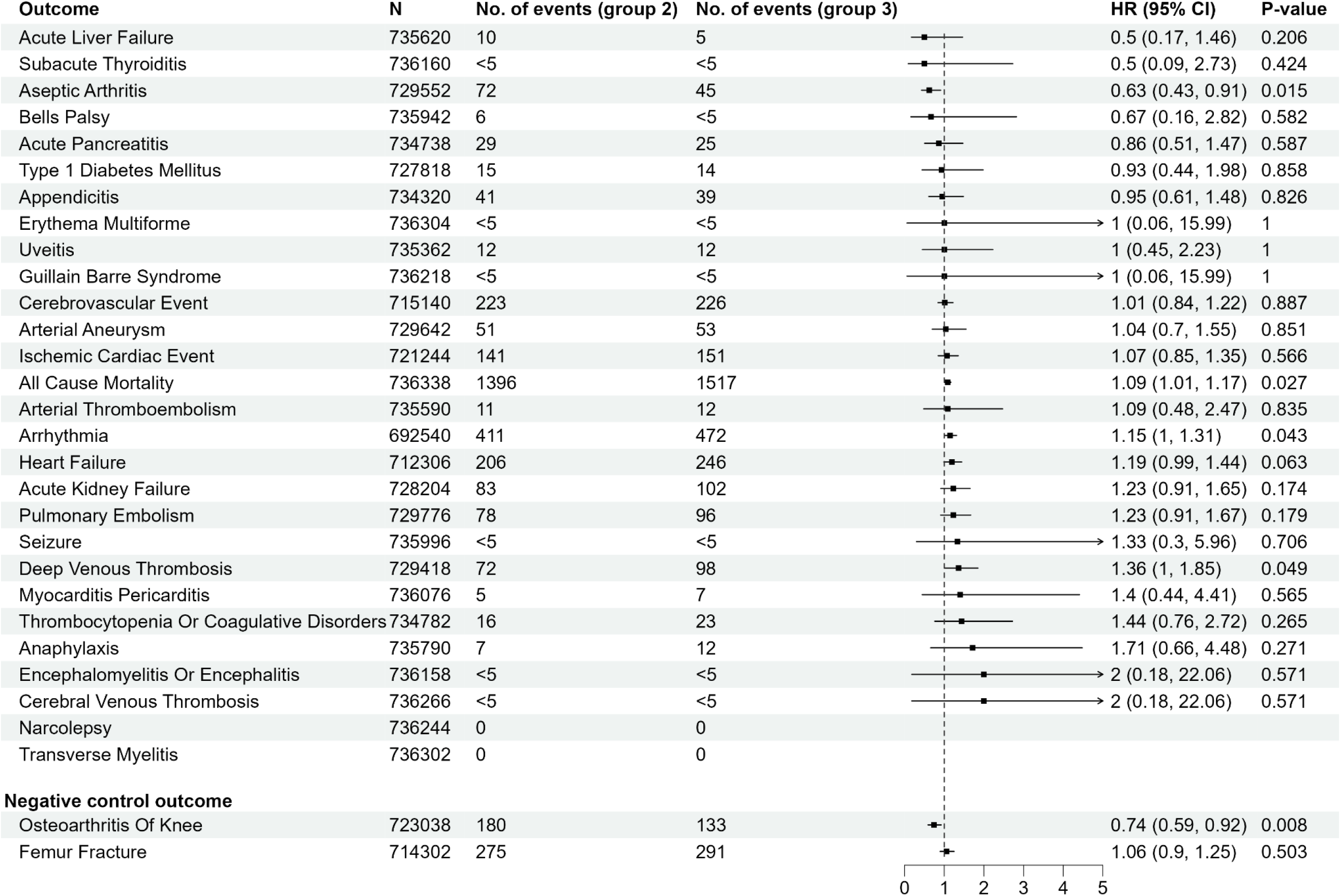
Main analysis comparing vaccine batch group 3 and vaccine batch group 2.

We conducted post-hoc sensitivity analyses of the association with arrythmia in both comparisons (figure 4). There was no consistent pattern between the two comparisons. In the group 3 vs group 1 comparison, the effect was largest among <40-year-olds (HR 5.00, 95% CI, 1.10-22.82), females (1.31, 1.02-1.68) and health-care personnel (2.09, 1.25-3.49), although confidence intervals were overlapping. In contrast, in the group 3 vs group 2 comparison, the effect was similar across age groups, between sexes and between priority groups, with largely overlapping confidence intervals.

**Figure 4.**
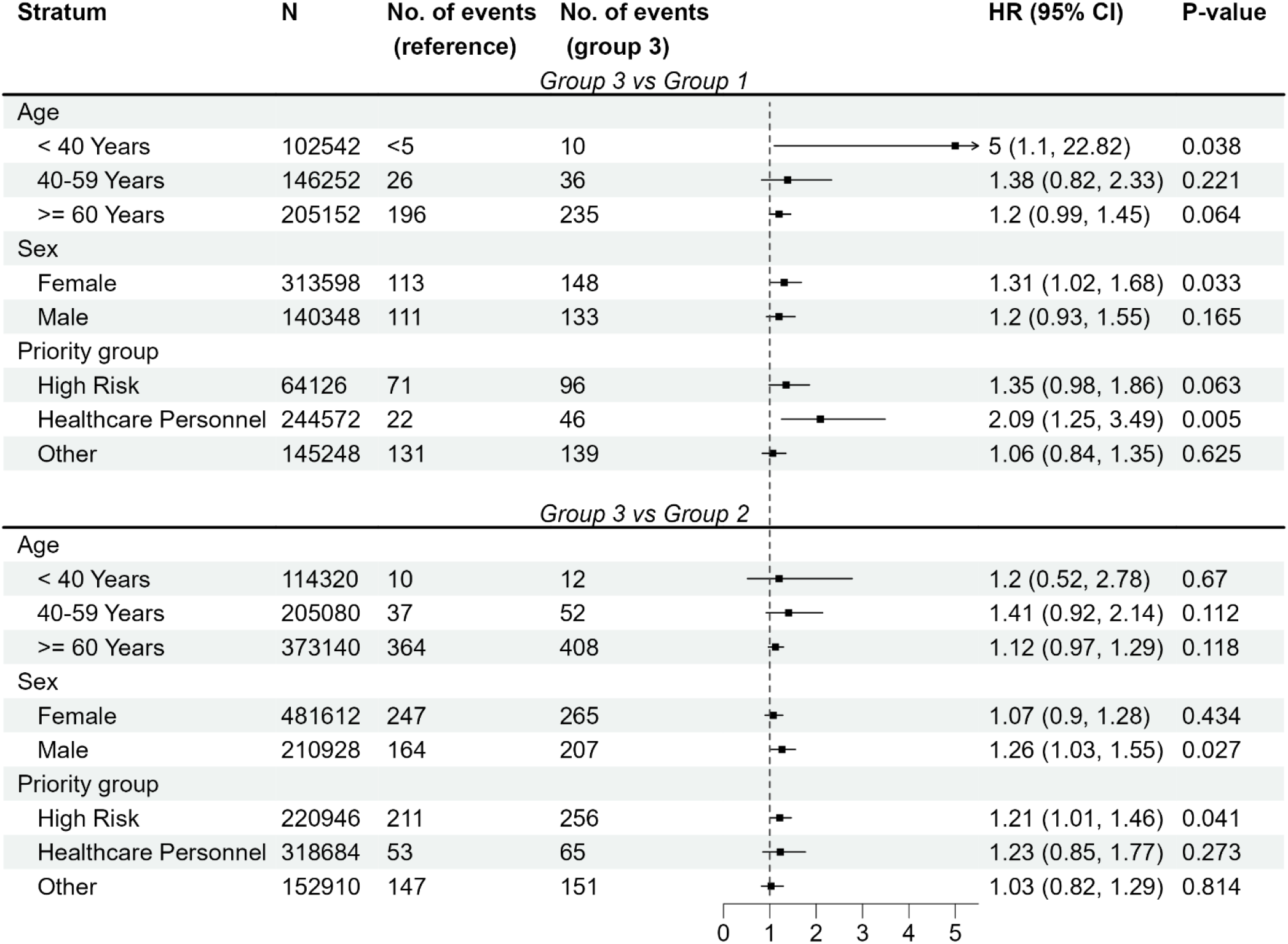
Stratified of hazard ratios of arrhythmia in both batch group comparisons.

## Discussion

Our findings suggest that the rates of serious adverse events, such as cerebrovascular and ischemic cardiovascular events, did not differ significantly across batch groups. The observations of slightly higher rates of arrhythmia in the first smaller batches of the vaccination roll-out were not robust to sensitivity analyses.

The results align well with the current evidence emphasizing the overall safety of the COVID-19 vaccines,[4] and they are in contrast to the Danish research letter which claimed large variations in batch-dependent safety.[1] The previous Danish study has a number of serious limitations.[7–9] The number of delivered vaccines do not equal administered vaccines within the study period. The study cut-off date 11 January 2022, means that a significant number of group 3 vaccinations would not even have been administered let alone have resulted in adverse events and reports. The study also compares reporting rates in individuals and time-periods that are not comparable. Group 3 comprised vulnerable elderly with multimorbidity and frontline personnel vaccinated at a time when the authorities recommended that all adverse events, including events such as sore shoulders and fever, were reported. Groups 1 and 2 comprised primary course- and booster vaccinations in the general population at a time when it was not recommended to report common and well-known adverse events.

Our study circumvents many of the weaknesses of the previous research letter. We use diagnostic endpoints instead of pharmacovigilance reports and utilises a matched design which compares individuals of the same age, sex and from the same vaccination priority group. However, our study also has a number of weaknesses. First, despite matching, residual confounding is still a possibility. Second, group 3 is more likely to be first doses, where group 2 comprises both first and second doses, and group 1 comprises booster doses. The reactogenicity of the vaccine may differ according to prior levels of immunity. Third and final, we are comparing vaccinations between different time periods. If there are strong calendar period trends in the study outcomes, we are not able to take this into account due to the almost perfect correlation between the batch groups and the periods in which they were administered. We believe this is reflected in the contrast of the all-cause mortality results between comparisons. The slightly increased rate observed in the group 3 vs group 2 comparison is a comparison between vaccinations administered at a median date of January 30, 2021 and vaccinations administered at a median date of June 26, 2021, i.e. a comparison between all-cause mortality in winter and summer. In contrast, no increased rate was observed in the group 3 vs group 1 comparison which is a comparison between vaccinations administered at a median date of January 30, 2021 and vaccinations administered at a median date of December 20, 2021, i.e. a comparison of all-cause mortality in two winter periods.

The batches included in this evaluation are not unique to Denmark, nor are the concerns about batch safety of the mRNA COVID-19 vaccines. Our results from the first nationwide cohort study of batch safety with individual-level data on vaccination and diagnoses provide reassurance that the safety of the BNT162b2 vaccine did not vary to any clinically relevant extent between batches used in Denmark between December 27, 2020 and to April 25, 2023. Currently, there is no compelling evidence to suggest otherwise.

## Data Availability

The data in this study is individual-level health information which is sensitive and cannot be shared by the authors.

## Ethical approval

The analyses were performed as surveillance activities as part of the advisory tasks of the governmental institution Statens Serum Institut (SSI) for the Danish Ministry of Health. SSI ‘s purpose is to monitor and fight the spread of disease in accordance with section 222 of the Danish Health Act. According to Danish law, national surveillance activities conducted by SSI do not require approval from an ethics committee. Both the Danish Governmental law firm and the compliance department of SSI have approved that the study is fully compliant with all legal, ethical, and IT-security requirements and there are no further approval procedures required for such studies.

## Author contributions

All authors conceptualised the study, interpreted the results, and critically reviewed the manuscript. AH drafted the manuscript, IBS carried out the statistical analyses. AH supervised the study. IBS had full access to all the data in the study and takes responsibility for the integrity of the data and the accuracy of the data analyses. AH is the guarantor. The corresponding author attests that all listed authors meet authorship criteria and that no others meeting the criteria have been omitted.

## References

1 Schmeling M, Manniche V, Hansen PR. Batch-dependent safety of the BNT162b2 mRNA COVID-19 vaccine. Eur J Clin Invest. 2023;53:e13998.

2 Vibeke Manniche – corona. https://vibekemanniche.dk/tag/corona/ (accessed 18 December 2023)

3 Grove Krause T, Jakobsen S, Haarh M, et al. The Danish vaccination register. Euro Surveill Bull Eur Sur Mal Transm Eur Commun Dis Bull. 2012;17:20155.

4 Andersson NW, Thiesson EM, Hansen JV, et al. Safety of BA.4-5 or BA.1 bivalent mRNA booster vaccines: nationwide cohort study. BMJ. 2023;382:e075015.

5 Lynge E, Sandegaard JL, Rebolj M. The Danish National Patient Register. Scand J Public Health. 2011;39:30–3.

6 Pedersen CB. The Danish Civil Registration System. Scand J Public Health. 2011;39:22–5.

7 Hviid A. Inaccurate representation of shipped vaccines as administered. Eur J Clin Invest. 2023;53:e14066.

8 del Saz BS. Batch-dependent safety of the BNT162b2 mRNA COVID-19 vaccine. Eur J Clin Invest. 2023;53:e14050.

9 Scott DAR, Renelle A, Niederer RL. Methodological and statistical considerations: Batch-dependent adverse effects of COVID-19 vaccines. Eur J Clin Invest. 2023;53:e14073.

